# The 10-year health impact, economic impact, and return on investment of the South African molecular diagnostics programme for HIV, Tuberculosis and SARS-CoV-2

**DOI:** 10.1101/2024.03.27.24304888

**Authors:** Brooke E. Nichols, Alexandra de Nooy, Naseem Cassim, Lucia Hans, Manuel Pedro da Silva, Kamy Chetty, Kyra H. Grantz, Alvin X. Han, Andrew N. Phillips, Lise Jamieson, Lesley E. Scott, Wendy S. Stevens

## Abstract

To ensure there is adequate investment into diagnostics, an understanding of the magnitude of impact and return on investment is necessary. We therefore sought to understand the health and economic impacts of the molecular diagnostic programme in South Africa, to deepen the understanding on the broad value of diagnostics and guide future healthcare investments. We calculated the 10-year (where data were available) total cost and DALYs averted associated with molecular diagnosis of molecular TB testing (2013-2022), HIV viral load monitoring (2013-2022), early infant diagnosis of HIV infection (2013-2022), and SARS-CoV-2 testing (2020-2022). We then calculated the economic value associated with those health gains and subsequent return on investment. Since the inception of the molecular diagnostics programme in South Africa, 3,035,782 DALYs have been averted as a direct consequence of this programme. This has generated an estimated $20.5 billion in economic value due to these health gains. The return on investment varied by specific diagnostic test (19.0 for tuberculosis, 1.4 for HIV viral load testing, 64.8 for early infant diagnosis of HIV, and 2.5 for SARS-CoV-2), for an average of 9.9 for the entire molecular diagnostics programme between 2013 and 2022- or $9.9 of value for each $1 invested. The molecular diagnostics programme in South Africa generated a significant amount of health gains and economic value associated with these health gains, and the return-on-investment rivals other high-impact public health interventions such as childhood vaccination. Consequently, the molecular diagnostics programme in South Africa is highly impactful, and will continue to be an excellent investment of South African public health expenditure.

## INTRODUCTION

Diagnostics play a vital role in ensuring quality health care at an individual level - through disease diagnosis and screening as well as condition and treatment monitoring- and at the global, national, or regional level through population disease surveillance (1). However, the importance of diagnostics is frequently overlooked, resulting in underfunded or under-resourced programmes (2). It is estimated that approximately 47% of the global population have limited (if any) access to diagnostics, with alarming disparity between the Global North and the Global South (2). Further, in comparison with other key healthcare elements, like medicine, investment in diagnostics has historically been limited (3). This is concerning given the direct connection between diagnosis and treatment- and has resulted in significant empiric treatment across the disease spectrum, resulting in over-, under-, or incorrect treatment (3). Additionally, although there is notable evidence of the substantial diagnostic gap in many low- and middle-income countries (LMICs) - particularly for key disease areas like HIV, tuberculosis (TB) and diabetes - there is little high-level literature which demonstrates the direct and long-term health and economic impacts of strong and well-distributed public health diagnostic programmes (4). This lack of evidence has limited the broad country-level investments in diagnostics needed to ensure quality health care for all.

In terms of diagnostic access, South Africa is a leader within the South African Development Community (SADC) region of sub-Saharan Africa. The country’s National Health Laboratory Service (NHLS) has a network of centralised molecular laboratory facilities and a daily specimen transportation and results delivery system. There is cover at all public healthcare facilities (PHCs) across the country, to provide diagnostic service support to ∼85% of the total population and 100% of the public sector (5–7). Further, the NHLS conducts millions of tests (both molecular and other) each year across a range of diseases, with nearly 107 million tests being performed in 2021-2022 (8). Of these, approximately 10 million (9.4%) were molecular tests performed by the NHLS that support the long-standing HIV and TB treatment programmes and more recently the severe acute respiratory syndrome coronavirus 2 (SARS-CoV-2) programme (8,9). These include HIV viral load testing, early infant diagnosis (EID) for HIV infection, tuberculosis diagnosis, as well as SARS-CoV-2 diagnosis. Over the last decade, molecular diagnostics have been introduced to each programme at different time points. In 2004, molecular diagnostics were introduced for viral load monitoring of people living with HIV on antiretroviral treatment. In 2011, the molecular diagnostics programme for TB was introduced, replacing the standard of care for TB diagnosis at the time-the less accurate smear microscopy. The molecular diagnosis programme expanded to include early infant diagnosis of HIV in 2013. All these molecular platforms were leveraged for the diagnosis of SARS-CoV-2 infection in 2020.

To ensure there is ongoing adequate and sustainable investment in diagnostics, an understanding of the magnitude of impact and return on investment is necessary. We, therefore, investigated the ten-year health and economic impacts of the public health molecular diagnostic programme in South Africa to deepen the understanding on the broad value of diagnostics to guide future healthcare investments.

## METHODS

### Health impact

The health impact, in terms of disability adjusted life years (DALYs), of each of the molecular diagnostic tools was calculated separately (Table 1). The health impact for TB, HIV viral load, early infant diagnosis of HIV, and SARS-CoV-2 molecular testing were all sourced from the literature or updated from existing models in the literature to understand both direct clinical benefit of diagnosis and the extended benefits that diagnostic testing or monitoring associated with a potential reduction in disease transmission can have. Impact can result from multiple tests of a single individual (e.g. a TB diagnosis and a viral load monitoring test), and these benefits are assumed to be additive.

**Table 1.**
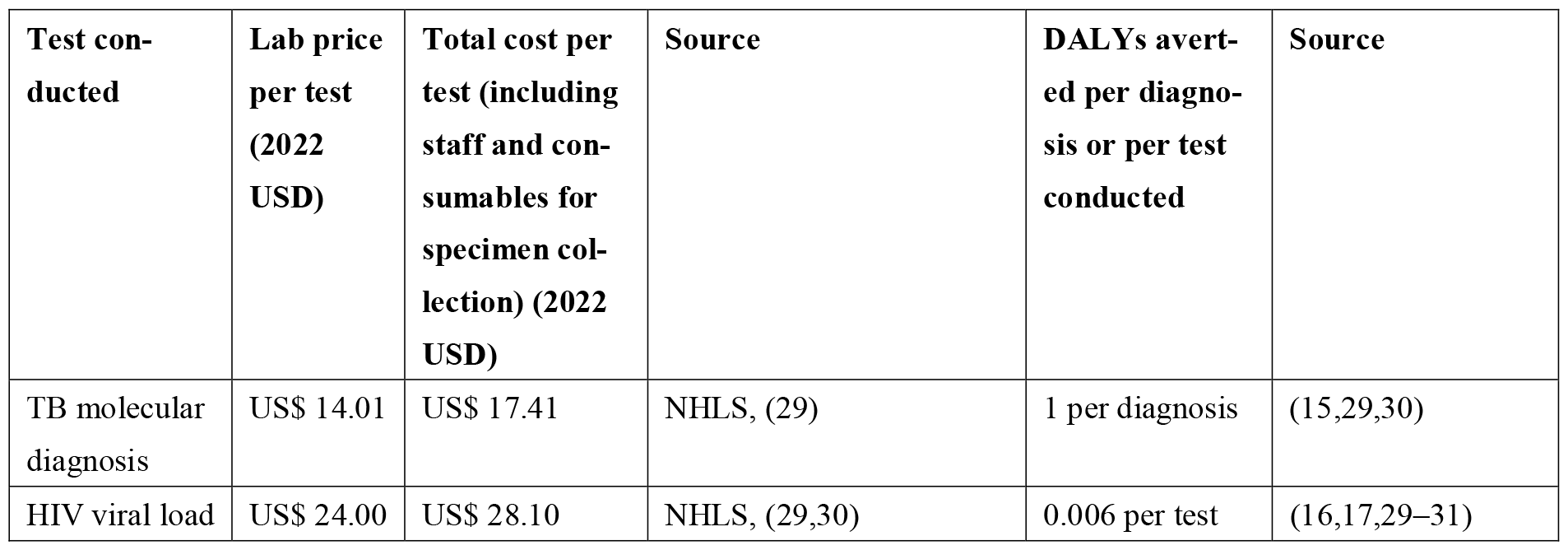
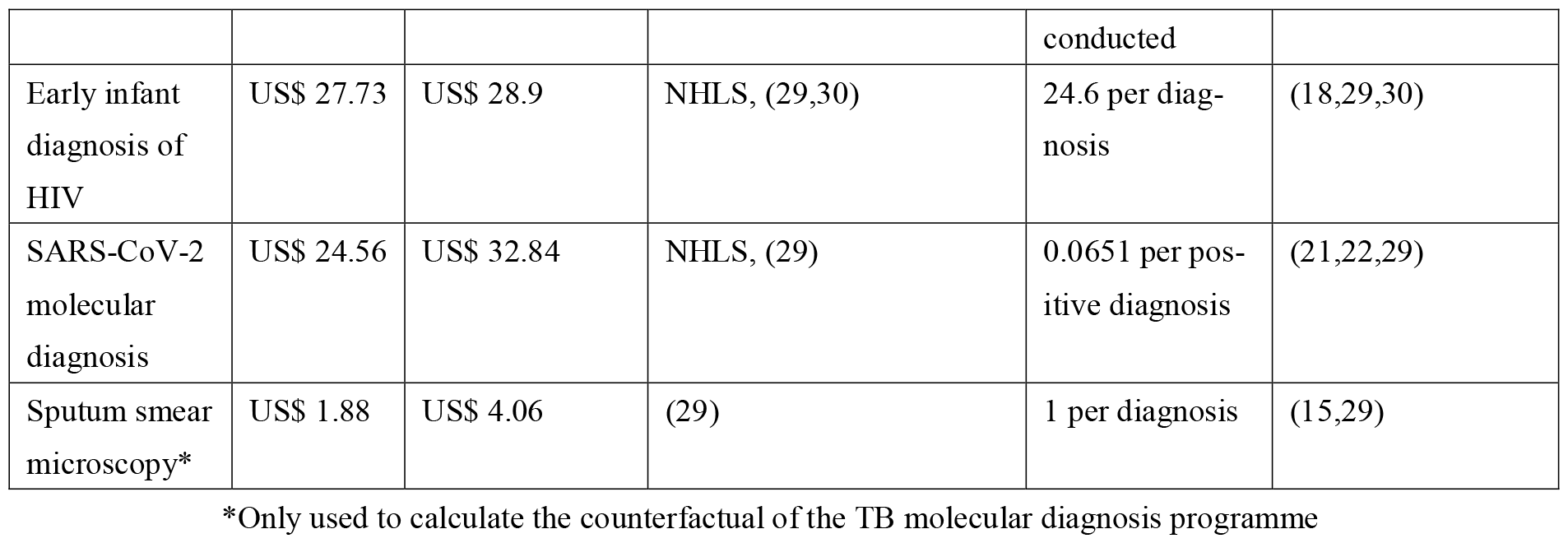
The cost per test and DALYs assumed per test or diagnosis as noted per disease area supported by the molecular diagnostics programme in South Africa.

**Table 2.** The return on investment of the molecular diagnostics programme in South Africa, as expressed by the total economic value of the health returns divided by the costs of conducting those tests across all years considered in this analysis.

| Test conducted | Economic value of health gains (2022 USD) | Total cost of implementation (2022 USD) | Return on investment |
| --- | --- | --- | --- |
| TB molecular diagnosis* | US\$ 6.4 billion | US\$ 337 million** | 19.0 |
| HIV viral load | US\$ 1.7 billion | US\$ 1.27 billion | 1.4 |
| Early infant diagnosis of HIV | US\$ 6.1 billion | US\$ 179 million** | 64.8 |
| SARS-CoV-2 molecular diagnosis | US\$ 0.6 billion | US\$ 273 million | 2.5 |
| Total | US\$ 20.5 billion | US\$ 2.1 billion | 9.9 |

\*Incremental benefit compared to a counterfactual of a smear microscopy programme
\*\*Including healthcare costs associated with false positive test results

#### Diagnosis of tuberculosis

The total number of TB tests conducted and number of positive diagnoses through the molecular diagnostics programme in South Africa was sourced from the literature(10). We compared the total net impact of the molecular TB programme as well as the incremental impact compared to if the sputum smear microscopy (SSM) programme had continued as it was in 2010. For the counterfactual SSM programme, we assumed that 1.329 million tests per year would be conducted from 2013-2022 (the same number of tests conducted in 2010 when the SSM diagnostic programme was at capacity)(11). We then assumed a constant 10% prevalence of TB among those tested, and a 68% test sensitivity, and a 98% test specificity of sputum smear microscopy (11). The sensitivity and specificity of Xpert MTB/Rif (12) was assumed to be 85% and 98% respectively, and the sensitivity and specificity of Xpert MTB/Rif Ultra (13), was assumed to be 91% and 99% respectively(14). For the total impact of the programme, we multiplied the number of DALYs averted per correct positive TB diagnosis (assumed to be 1 DALY per diagnosis (15)) by the number of correct TB diagnoses reported over time(10) (adjusting for test specificity). The benefits of the molecular programme are results of the improved accuracy of molecular diagnostics compared to smear microscopy for TB diagnosis.

#### HIV viral load monitoring

The HIV Synthesis model, an individual-based stochastic simulation model that includes HIV transmission, progression, HIV-related treatment, and prevention interventions, was used to estimate the DALYs associated with a viral load test (16,17). Two scenarios were run to tease out the incremental benefit of viral load testing: 1) no viral load testing for three years, followed by fully implemented viral load testing, and 2) fully implemented viral load testing over 10 years. The difference in DALYs between the two scenarios provides an indication of the DALYs averted because of viral load testing alone. To account for stochasticity, the model was run 300 times to produce an estimated average of 1250 DALYs averted per 3 months, in the context of an adult population of 10 million with 207,000 viral load tests done per 3 months compared to when no viral load tests were done. This resulted in 0.006 DALYs averted for each viral load conducted.

We then multiplied the number of viral load tests by the number of DALYs averted per viral load conducted across the lifespan of the NHLS molecular diagnostics programme. Given that we calculated the impact per viral load test conducted (rather than per elevated viral load result), specific test accuracy parameters were not taken into account.

#### HIV early infant diagnosis (EID)

A model of quality assurance systems for HIV EID estimated the DALYs associated with false negative test results with and without these systems for five countries(18). This study indicated that the DALYs associated with an undiagnosed case of HIV among infants was 24.6 in South Africa. We then multiplied the number of DALYs averted by the number of infants correctly diagnosed with HIV by year as reported in the literature(19) (adjusting for test specificity). The sensitivity of HIV EID is assumed to be 99.3% and specificity 99.5% (20), and a second test is required for confirmation.

#### Diagnosis of SARS-CoV-2

We used results from the Propelling Action for Testing and Treating (PATAT) simulation model of SARS-CoV-2 transmission to estimate the DALYs averted per SARS-CoV-2 diagnostic test used in countries most similar to South Africa in terms of population demography and household size (21,22).The impact of testing was primarily due to the reduction in the number of onward infections following isolation of positive cases, rather than any specific clinical impact. Individuals were assumed to reduce their number of community contacts by 50% on average after receiving a positive diagnosis but were not assumed to be able to effectively isolate from household members. Given that the relative impact of testing for SARS-CoV-2 is dependent on several parameters, we averaged the impact across effective reproductive numbers of SARS-CoV-2 from 1.2-2.0, a testing rate of 100 tests per 100,000 per year, and minimal vaccine coverage. This resulted in 0.0651 DALYs averted per correct positive test result (adjusted for test specificity). The sensitivity of molecular SARS-CoV-2 testing was assumed to be 99% and specificity 97% (23).

### Economic impact of health gains and return on investment (ROI)

The total number of DALYs per health area were calculated per year and multiplied by the GDP per capita per respective year to calculate the economic impact of the molecular diagnostics programme. Results are reported for each diagnostic test, and for the entire programme (Table 1). No DALYs are averted for false positive results at the end of a diagnostic algorithm, calculated using the average expected incidence of infection for each health area and the specificity of the test or test algorithm. All health benefits and related economic gains are expressed in terms of net present value of these gains. The health benefits (DALYs averted) of a TB molecular diagnostic are expected to occur over a lifetime after the average age of active TB disease, the benefits of HIV viral load testing were calculated over a ten-year period (discounted at 3% annually), early infant diagnosis over a lifetime (discounted at 3% annually), and SARS-CoV-2 testing over 1 year (as such, not discounted).

Return on investment was calculated as the total economic impact per diagnostic tool divided by the total cost of all diagnostic tests required to generate that impact (including both “positive” and “negative” tests). The total number of tests per programme, across the respective time periods considered, were taken from the literature for TB, HIV viral load, SARS-CoV-2, and EID (10,19,24–26). To calculate the number of HIV EID tests conducted on average for 2020-2022, we used the number of live births to women living with HIV in 2021 (∼275 527), the EID testing algorithm in South Africa and the coverage of testing at each part of the algorithm (averaged between NICD and DHIS sources). Given the relative overcounting of infants diagnosed with HIV reported in the literature(19), we also adjusted the numbers of positive infants reported in 2013-2019 down to reflect this overcounting. We also adjusted for the neonatal mortality rate in South Africa for number of infants expected to be tested at 10 weeks, and the number of infants testing positive who would not need to be retested (27).

The total costs were calculated as the 2022 test price per test in South African Rands (ZAR), converted to United States Dollar (USD) using average 2022 conversion rate (16.37 ZAR to 1USD (28)), and multiplying each respective test cost by the total number of tests of that type conducted per year since the inception of the molecular diagnostic programme. “Deadweight” costs have been removed from the ROI analysis for TB diagnosis by using the appropriate incremental gains of the molecular programme (e.g. removing the costs associated with the counterfactual of keeping the sputum smear microscopy programme running at the same scale as when the molecular programme had been introduced). The additional costs incurred associated with false positive test results for TB and EID were calculated using prevalence of infection and test specificity of the testing algorithm. These additional costs included those for unnecessary treatment of drug sensitive TB disease, US$ 112.70 (29), and unnecessary lifetime ART treatment cost for an misdiagnosed with HIV, US$ 6,115 (discounted at 3% per year, assuming a life expectancy of 63.8 years) (18).

### Ethics

Solely aggregate data from previously published manuscripts was used for this analysis. (10,19,24,25). The data are available *open access* in each publication. Any missing data points were imputed rather than extracted from raw data.

## RESULTS

### Health Impact

Since the inception of the molecular diagnostics programme in South Africa, 3,035,782 DALYs have been averted as a direct consequence of this programme. This includes 961,499 incremental DALYs averted due to the diagnosis of TB (2013-2022) (incremental to a continued smear microscopy programme, adjusting for false positive test results), 272,009 averted due to HIV viral load tests (2013-2022), 1,700,208 due to early infant diagnosis (2013-2022), and 102,066 DALYs averted due to molecular SARS-CoV-2 testing (2020-2022). Figure one highlights the relative impact per year that each of the diagnostics has demonstrated.

### Economic impact of health gains

The total economic value of the molecular diagnostics programme due to health gains over the ten-year period was US$ 20.5 billion. This includes US$ 6.4 billion in economic value associated with improved health related to TB diagnosis (2013-2022), US$ 1.8 billion for HIV viral load testing (2013-2022), US$ 11.6 billion for HIV early infant diagnosis (2013-2022), and $0.7 billion for SARS-CoV-2 testing (2020-2022) (Figure 2).

**Figure 1.**
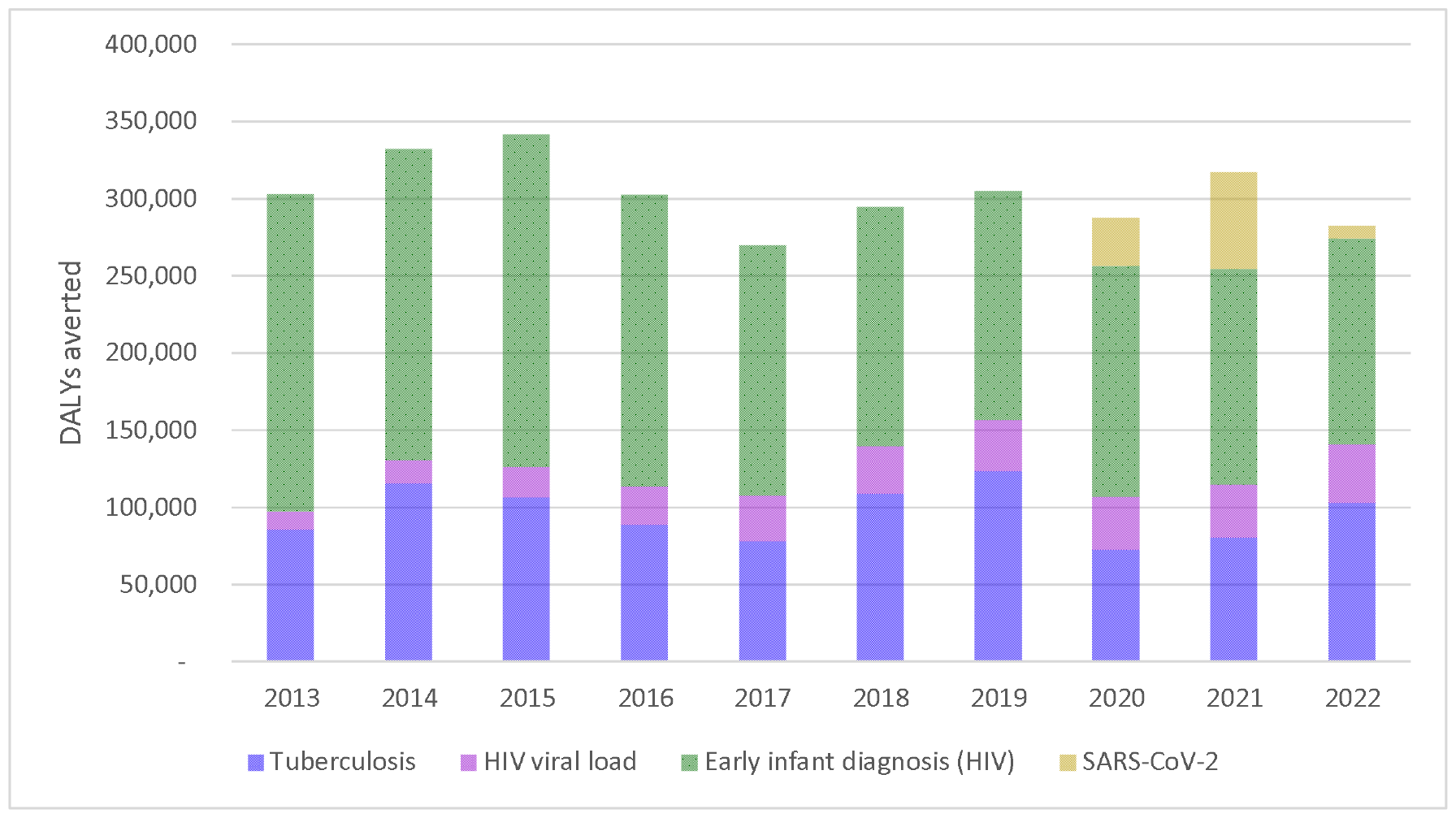
The annual DALYs averted of South Africa’s public sector molecular diagnostics programme for HIV, TB and SARS-CoV-2 from 2013-2022

**Figure 2.**
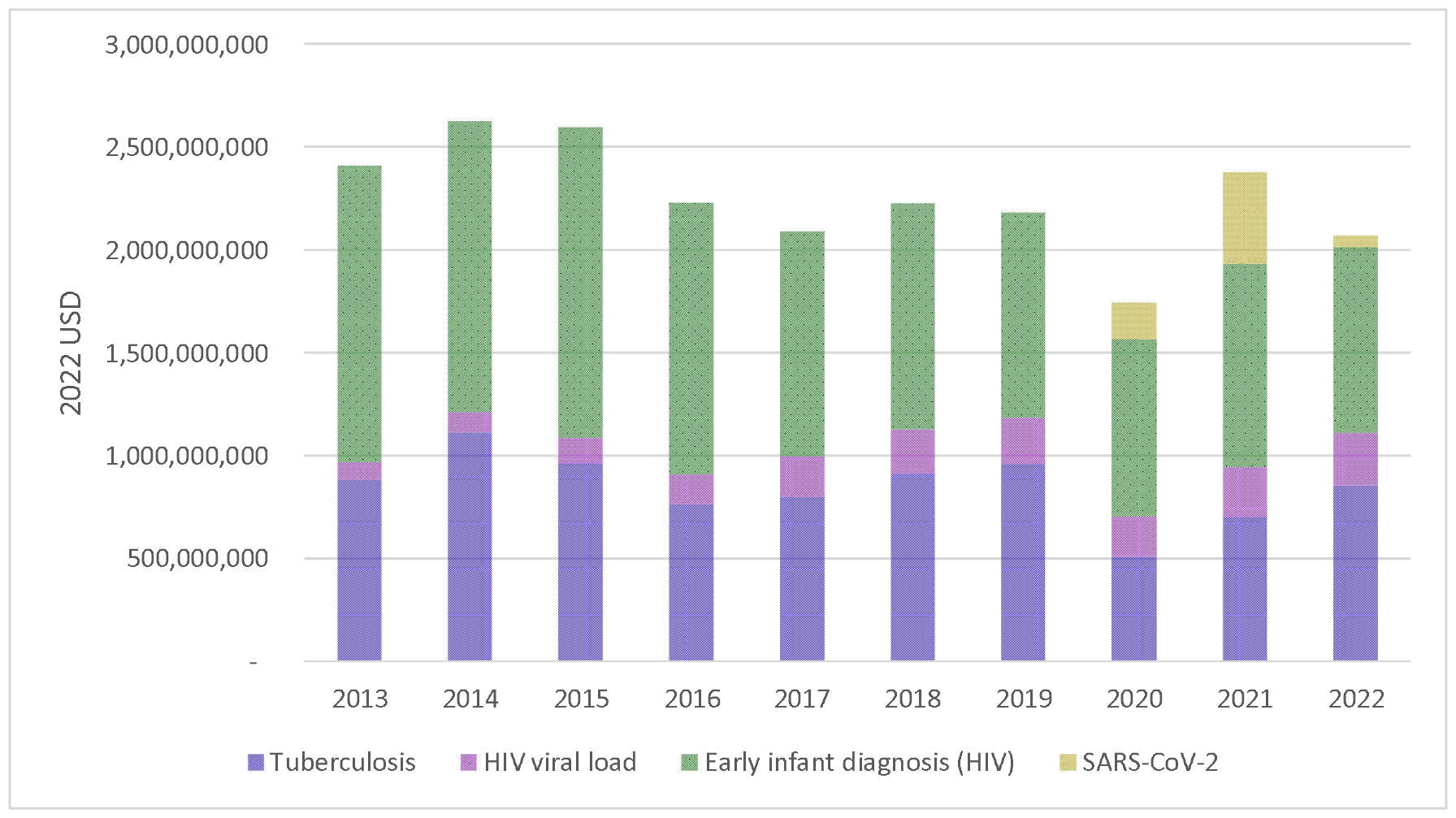
Annual economic value (in 2022 USD) associated with the health gains of the molecular diagnostics programme for HIV, TB and SARS-CoV-2 from 2013-2022

### Return on investment

The molecular TB diagnostics programme has cost an estimated US$ 418 million over 10 years (for all TB diagnostics conducted plus costs associated with false positive test results), while our counterfactual was assumed to cost $81 million over the same period (for diagnostics plus costs associated with false positive results), for an incremental cost of USD$337 million of the molecular TB diagnostics programme. The total incremental economic value of health gains associated with TB molecular diagnosis divided by the total incremental cost of the molecular TB diagnostics programme resulted in a return on investment of 19.0- or US$ 19.0 of economic value for each US$ 1 invested in diagnosis. The return on investment for HIV viral load was 1.4, for HIV early infant diagnosis was 64.8, and for SARS-CoV-2 diagnosis was 2.5. Averaged across the whole molecular diagnostics programme, the average return on investment was 9.9 - or US$ 9.9 of economic gain related to improved health for each US$ 1 invested in diagnostics.

## DISCUSSION

Generally, molecular testing has multiple roles in disease management, including diagnosis, monitoring response to treatment and guiding decisions on appropriate management (e.g. indications for HIV drug resistance tests) (9). Further, as compared to other diagnostic counterparts (like TB culture), molecular tests are frequently able to return accurate results more rapidly, and hence allow for action to be taken at an earlier stage (9). Thus, molecular testing, if well-implemented, has the potential to greatly improve patient outcomes. This has been shown by the public sector molecular diagnostics programme in South Africa that generated a significant amount of health gains and economic value associated with these health gains. The return on investment was ≥1.4 for all individual tests, as high as 64.8 and 9.9 on average-suggesting excellent value for money across all molecular tests. These levels of return on investments for some tests rival childhood vaccination programmes (ROI 16)-often considered one of the most valuable public health interventions (32,33). While the ROI value for each of the individual types of molecular tests varied greatly, each still resulted in a positive impact on the South African healthcare system given that all tests had at least a full ROI.

Comparing to other public health tools such as vaccines and therapeutics, for each specific disease area is useful to understand the relative role of diagnostics as part of a public health programme. For SARS-CoV-2, the ROI of vaccination has not been estimated for South Africa, but in New York City was estimated to be 10 to 1(34). If adjusting this estimate for the GDP per capita in South Africa, and the smaller proportion of people at risk for severe disease and death in South Africa (due to demographic differences), the ROI for SARS-CoV-2 vaccines in South Africa might approach the value of diagnostics in South Africa. ROIs for specific therapeutics for SARS-CoV-2 in South Africa have not been calculated. Therapeutics such as paxlovid, however, have not been available in South Africa. For both TB and HIV, the return on investment for investments into vaccines has been effectively 0 (or negative) given that there are no vaccines available for HIV, and no effective vaccines for adults for TB (with the BCG vaccine for TB protective for <5-year-olds only). For HIV, the UNAIDS “Fast-Track” strategy which focuses on investments in expanding HIV service coverage (as opposed to maintaining constant coverage) estimates an approximate return on investment of 6.46 to 1 for Southern Africa (35,36). This contextualizes that viral load testing is complementary to a broader package of HIV-related services, with an ROI of 2, and that early infant diagnosis remains to have an extraordinarily high ROI. With respect to TB, there also exists a set of services which together have resulted in an estimated drop of more than 50% in the incidence rate between 2011 and 2022 (37). While it is difficult to quantify exactly how much each particular service (e.g. the increase in TB diagnostic capacity and accessibility, increased TB treatment options and the impact of improved HIV services) has contributed towards reduced incidence, the introduction of Xpert MTB/Rif and Ultra and the scale-up of the molecular testing programme has clearly contributed. Similarly in terms of investment, while there is no specific literature available on the return-on-investment of TB diagnostics, studies have shown that TB prevention and care yields a ROI of 43-implicitly including diagnosis-whereas we have estimated the ROI of diagnosis aspect to prevention and care to be 19.0 (38).

These values represent one consistent methodological way to calculate impact, economic value, and ROI. There are several changes in the underlying assumptions that might influence the direction of these values in both directions. Firstly, the average GDP per capita in South Africa may not reflect the burden of disease where the health gains occurred for each respective disease (e.g. slightly lower burden of disease among higher household social economic index score for tuberculosis infection) (39). If including these differences, the economic gains might be slightly overstated where the benefits occurred amongst individuals with lower annual GDP. We did not, however, want to weight the economic value of health differently among the South African population. In the other direction, within the scope of this analysis, we have only calculated the economic value of the health gains associated with molecular diagnostics rather than the *total* economic benefit. The total economic benefit likely far exceeds what we have estimated here-for example, the benefits associated with being able to return to work after a negative SARS-CoV-2 test during the pandemic or benefits associated with the reduced frequency of healthcare clinic visits required after a suppressed viral load test-reducing costs to individuals on antiretroviral treatment and possibly increasing economic activity due to fewer days spent seeking care. Estimating the total value was outside the scope of this analysis.

There were several limitations to our analysis. First, we did not include the value related to drug susceptibility testing (DST) for tuberculosis given the paucity of literature on the DALYs associated with TB resistance testing. If we had included that, the likely value and impact of the molecular diagnostic programme would have only increased, especially since the diagnosis of rifampicin resistant TB is provided simultaneously with the molecular diagnosis of TB. Second, for the incremental impact of the TB molecular diagnostics programme, we assumed that the size of the smear microscopy programme would have held constant in size from 2010 onwards. An expansion of this programme would have generated more health effects and related economic value of health gains, resulting in a diminished incremental benefit of the molecular programme. We made this assumption, however, given that the microscopy programme was already at capacity in 2010. Third, for the HIV viral load DALY calculations, it was assumed that individuals would be on a dolutegravir-based first line antiretroviral treatment regimen. The DALYs per viral load would have likely been greater in pre-dolutegravir era, given that previous regimens were slightly less effective. This would have resulted in a slight underestimate of the benefit of viral load testing before the introduction of dolutegravir-based first-line antiretroviral treatment regimens-but do reflect the likely benefit moving forward. Fourth, while we have presented here the net present benefits of the molecular diagnostics programme, the time horizon of the economic benefit associated with health gains differ between diseases. Finally, we did not consider the impact of potential additional costs incurred as a result of test sensitivity given the uncertainty around the costs that may occur as a consequence (e.g. false negative tests). Individuals who receive a false negative result may need to seek care again (or multiple times) to receive a correct diagnosis, incurring additional costs (most likely with TB testing, less so for SARS-CoV-2 given the short duration of infection). These individuals may also transmit infection if they are diagnosed negative (for TB, SARS-CoV-2, possibly for viral load if individuals think they are suppressed; not likely for EID). It is unlikely, however, that these tests will influence our results significantly, and the relative test sensitivity of molecular diagnostics is relatively high compared to other available diagnostics.

## CONCLUSIONS

In conclusion, the molecular diagnostics programme in South Africa is highly impactful, and will continue to be an excellent investment of South African health expenditure. The value and return on investment of these diagnostic tools is in line with other highly impactful public health interventions and should continue to be valued as such. Into the future, molecular diagnostics are likely to play a more significant role in the monitoring and control of existing diseases (other than those described here) and, as seen for SARS-CoV-2, may be the first point of call for the diagnosis of and surveillance of future pandemics given their relative ease of development and implementation into existing systems.

## Data Availability

The data are available open access in each relevant publication. Any missing data points were imputed rather than from raw data.

## Author Contributions

Conceptualization, BEN, LES, WSS; methodology, BEN; formal analysis, BEN, KHG, AXH, ANP, LJ; data curation, AdN, NC, LH, MPdS, KC, LES, WSS.; writing—original draft preparation, BEN AdN; writing—review and editing, (NC, LH, MPdS, KC, KHG, AXH, ANP, LJ, LES, WSS); All authors have read and agreed to the published version of the manuscript.

## Institutional Review Board Statement

Not applicable

## Informed Consent Statement

Not applicable

## Data Availability Statement

The data are available *open access* in each relevant publication. Any missing data points were imputed rather than from raw data.

